# Low vitamin K status predicts mortality in a cohort of 138 hospitalized patients with COVID-19

**DOI:** 10.1101/2020.12.21.20248613

**Authors:** A Linneberg, FB Kampmann, SB Israelsen, LR Andersen, HL Jørgensen, H Sandholt, NR Jørgensen, SM Thysen, T Benfield

## Abstract

It has recently been hypothesised that Vitamin K could play a role in COVID-19. We aimed to test the hypothesis that low vitamin K status is a common characteristic of patients hospitalized with COVID-19 compared to population controls; and that low vitamin K status predicts mortality in COVID-19 patients. In a cohort of 138 COVID-19 patients and 140 population controls, we measured plasma dephosphorylated-uncarboxylated Matrix Gla Protein (dp-ucMGP), which reflects the functional Vitamin K status in peripheral tissue. Fourty-three patients died within 90-days from admission. In patients, levels of dp-ucMGP differed significantly between survivors (mean 877; 95% CI: 778; 995) and non-survivors (mean 1445; 95% CI: 1148; 1820). Furthermore, levels of dp-ucMGP (pmol/L) were considerably higher in patients (mean 1022; 95% CI: 912; 1151) compared to controls (mean 509; 95% CI: 485; 540). Cox regression survival analysis showed that increasing levels of dp-ucMGP (reflecting low Vitamin K status) were associated with higher mortality risk (sex- and age-adjusted hazard ratio per doubling of dp-ucMGP was 1.50, 95% CI: 1.03; 2.18). In conclusion, we found that low Vitamin K status predicted mortality in patients with COVID-19 supporting a potential role of Vitamin K in COVID-19.

## 1. Introduction

Vitamin K is an essential vitamin for activation of blood clotting factors and several other proteins indicating that vitamin K has effects on coagulation, bone formation and inhibition of calcification in arteries. Vitamin K serves as a co-factor for the enzyme γ-glutamate carboxylase that converts glutamate residues into γ-carboxyglutamate (Gla). These Gla-residues serve as calcium-binding groups, which are essential for the activity of all Gla-containing proteins. Vitamin K status can be objectively assessed in two different ways: A) by measuring the Vitamin K concentration in plasma or B) by determining the amount of uncarboxylated vitamin K-dependent proteins. The first method reflects a snapshot of recent vitamin K intake, is sensitive to triglyceride concentrations, and gives limited information about the vitamin K utilization in tissue. In contrast, measurement of the inactive (uncarboxylated) form of Matrix Gla Protein (MGP) in the blood can be used as a biomarker of the functional vitamin K status in peripheral tissues. Thus, the dephosphorylated-uncarboxylated iso-form of MGP (dp-ucMGP) reflects the functional Vitamin K status and is considered the gold standard for measuring Vitamin K status in peripheral tissue.^1,2^

Corona virus disease 2019 (COVID-19) is a transmittable viral infection caused by Severe Acute Respiratory Syndrome Corona Virus-2 (SARS-CoV-2). COVID-19 presents very differently in patients; many patients experience mild symptoms, while other patients develop severe disease, including respiratory distress syndrome with high risk of death.^3^ A recently published study found significantly higher levels of dp-ucMGP (reflecting lower vitamin K status) in hospitalized COVID-19 patients compared to controls suggesting a role of vitamin K in COVID-19.^4^ Among the COVID-19 patients low vitamin K status was associated with a poor outcome of disease (need of invasive ventilation or death). Furthermore, low vitamin K status was associated with increased blood levels of desmosine, a biomarker of degradation of elastic fibres in the lung tissue,^5^ suggesting that low vitamin K status could increase the rate of degradation of elastic fibres during severe COVID-19. The authors hypothesised that increased degradation of elastic fibres in the lungs could be due to lack of activated MGP, which is known to protect extracellular matrix proteins such as elastic fibres from calcification and subsequent degradation^6^. MGP is the strongest known inhibitor of tissue calcification in the arterial vessel wall and thus prevents arterial calcification.^7^ MGP is also highly expressed in the lungs.^8,9^ Degradation of elastic fibres in the lungs stimulates calcification of elastic fibres.^10^ A rising calcium content of the extracellular matrix stimulates the local synthesis of MGP to prevent calcification of the elastic fibres.^6^ However, MGP is synthesised as dp-ucMGP, and needs activation by vitamin K-dependent carboxylation to be able to protect elastic fibres in the extracellular matrix from calcification. These processes could create a, or exacerbate a pre-morbid, vitamin K deficit during severe disease and increased demand of Vitamin K.

Aside from the carboxylation of prothrombotic proteins, vitamin K is also essential for carboxylation of antithrombotic proteins (e.g. Proteins S, Protein C)^11^. During a state of Vitamin K deficiency, e.g. increased vitamin K use during acute illness, vitamin K is primarily used for carboxylation of the prothrombotic coagulation factors in the liver and to a lesser extent for carboxylation of the extrahepatic vitamin K dependent proteins including MGP and the antithrombotic Protein S^4^. It has been hypothesised that this could induce a prothrombotic state with increased blood clotting in peripheral tissues as has been seen in COVID-19 patients.^9^

We aimed to test the hypothesis that low vitamin K status is a common characteristic of hospitalized patients with COVID-19 compared to population controls; and that low vitamin K status predicts mortality in hospitalized COVID-19 patients.

## 2. Material and Methods

### 2.1. The Amager Hvidovre Hospital COVID-19 cohort

Characteristics of the Amager Hvidovre Hospital (Hvidovre) COVID-19 cohort have previously been described.^12^ Briefly, this retrospective case series included adults 18 years of age or older with a new-onset pulmonary infiltrate and confirmed SARS-CoV-2 infection who were consecutively admitted between 10 March and 23 April 2020 at a 700-bed university-affiliated hospital in Copenhagen, Denmark. Cases were confirmed through reverse-transcriptase-polymerase-chain-reaction assays performed on an oropharyngeal swab or a lower respiratory tract specimen. Data including patient characteristics, vital parameters and laboratory measurements were transferred from electronic health records. The study was approved by the Danish Patient Safety Authority (record no. 31-1521-309) and the Regional Data Protection Center (record no. P-2020-492). A blood sample was drawn within 4 days from admission. In case more than one sample was drawn from the same patient, the dp-ucMGP measurement in the first sample was used in the statistical analyses. EDTA plasma was separated by centrifugation and stored at minus 80 °C. Measurements of dp-ucMGP in stored plasma samples from the biobank were approved by the Ethical Committee of the Capital Region of Denmark (record no. H-20047597).

### 2.2. General population controls

The population based Health2016^13^ study was performed at the Center for Clinical Research and Prevention (CCRP). A total of 4497 randomly selected persons living in 11 municipalities in the Western part of Copenhagen, covering part of the catchment area of Amager Hvidovre Hospital, were invited to a health examination and 1251 (28%) participated. All participants underwent a health examination and completed a questionnaire about lifestyle and health. Measurements of dp-ucMGP were performed in 491 consecutive participants (aged 19–71 years) examined between 30 May 2017 and 21 December 2017.^2^ For the present study participants aged 60 years or higher were used as population controls (n=141) to obtain an age distribution reasonably similar to that of the COVID-19 cohort. One participant taking Vitamin K antagonists was excluded. The Health2016 study was approved by the Ethics Committee of the Capital Region of Denmark (record no. H-15017277).

### 2.3. Measurements of Vitamin K status in plasma

In both patients and controls, biochemical determination of vitamin K status was performed by using the IDS-iSYS InaKtif MGP assay (Immunodiagnostic systems, plc, Tyne and Wear, UK) performed at the Department of Clinical Biochemistry, Rigshospitalet, Glostrup, Denmark.^2^ The InaKtif MGP assay is an in vitro diagnostic test intended for the quantitative determination of dp-ucMGP in human plasma on the IDS-iSYS Multi-Discipline Automated System. Control specimens had average values of 900 pmol/L, 4100 pmol/L, and 7050 pmol/L with intra-assay CV (coefficient of variation) of 3.9%, 0.9%, and 0.9% respectively. Inter-assay CVs were 3.2%, 2.8%, and 1.6% respectively. Since the reportable range is 300-12.000 pmol/L, dp-ucMGP values <300 pmol/L were fixed as 299 pmol/L.

### Statistical analyses

Dp-ucMGP was used as a continuous variable or categorical variable in three categories (lowest quartile, the two middle quartiles, and highest quartile of dp-ucMGP). Dp-ucMGP values were log-transformed (log2) in the analyses and back-transformed when presented. Baseline characteristics were reported as frequencies with percentages, means with SD or medians with interquartile range (IQR). Comparison between COVID-19 patient groups (non-survivor patients vs. survivor patients) was performed using Χ^2^-tests, Fisher’s-exact test, t-test, Mann-Whitney *U* or Monte Carlo simulation test as appropriate. Kaplan-Maier survival plots were drawn for each of the three dp-ucMGP categories and compared by the log-rank test. Hazard ratios (HRs) with 95% confidence intervals (CIs) were estimated using Cox proportional hazards models with time from blood sampling as underlying time scale and a doubling of dp-ucMGP as exposure. Patients entered the analysis at the time of blood sampling and was followed until death or 90 days after blood admission. Thus, the HR estimate reflects the relative risk for death within 90 days per one doubling of dp-ucMGP. The regression survival analyses were further adjusted for sex and age. A P-value < 0.05 was considered significant. Data analysis were performed using R software version 4.0.2 (R Foundation for Statistical Computing, Vienna, Austria).

## 3. Results

Dp-ucMGP was measured in a total of 138 COVID-19 patients. Thirty-six and 43 died within 30 and 90 days from inclusion, respectively. Table 1 shows characteristics of the COVID-19 patient cohort stratified by 90-day survival status. Among the patients, both 90-day mortality and 30-day mortality (data not shown) were significantly associated with high age, hypertension, cardiovascular disease, and increased levels of dp-ucMGP. Furthermore, levels of dp-ucMGP was higher among COVID-19 patients compared to the population controls (Table 1). The Cox regression analysis showed that the mortality risk was significantly higher in patients with increased levels of dp-ucMGP (unadjusted HR per doubling of dp-ucMGP was 1.94, 95% CI: 1.42; 2.67). The HR estimate attenuated after adjusting for sex and age (sex and age adjusted HR per doubling of dp-ucMGP was 1.50, 95% CI: 1.03; 2.18). When ending follow-up at 30 days from blood drawing, the mortality risk was 1.91 (unadjusted HR, 95% CI: 1.38; 2.68) and 1.45 (sex and age adjusted HR, 95% CI: 0.97; 2.15). A Kaplan-Meier plot of cumulated risk of dying versus time from blood drawing stratified by dp-ucMGP categories (lowest quartile, middle quartiles, and highest quartile) is shown in Figure 1. The log-rank test for comparison of the three dp-ucMGP categories was highly statistically significant (P<0.0001)

**Table 1:**
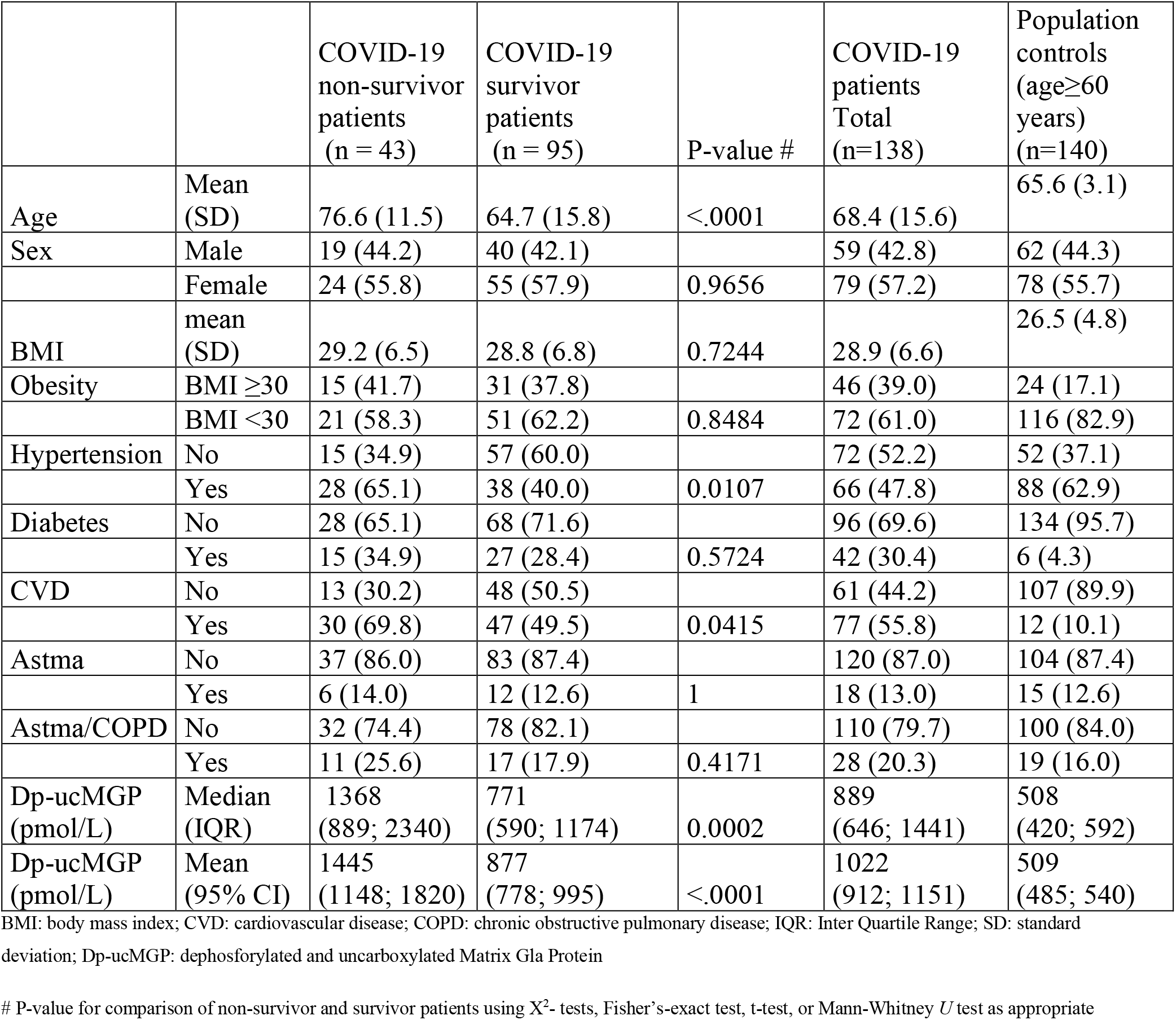
Characteristics of the COVID-19 patient cohort survivors and non-survivors, and general population controls

|  |  | COVID-19<br>non-survivor<br>patients<br>(n = 43) | COVID-19<br>survivor<br>patients<br>(n = 95) | P-value # | COVID-19<br>patients<br>Total<br>(n=138) | Population<br>controls<br>(age≥60<br>years)<br>(n=140) |
| --- | --- | --- | --- | --- | --- | --- |
| Age | Mean<br>(SD) | 76.6 (11.5) | 64.7 (15.8) | <.0001 | 68.4 (15.6) | 65.6 (3.1) |
| Sex | Male | 19 (44.2) | 40 (42.1) |  | 59 (42.8) | 62 (44.3) |
|  | Female | 24 (55.8) | 55 (57.9) | 0.9656 | 79 (57.2) | 78 (55.7) |
| BMI | mean<br>(SD) | 29.2 (6.5) | 28.8 (6.8) | 0.7244 | 28.9 (6.6) | 26.5 (4.8) |
| Obesity | BMI ≥30 | 15 (41.7) | 31 (37.8) |  | 46 (39.0) | 24 (17.1) |
|  | BMI <30 | 21 (58.3) | 51 (62.2) | 0.8484 | 72 (61.0) | 116 (82.9) |
| Hypertension | No | 15 (34.9) | 57 (60.0) |  | 72 (52.2) | 52 (37.1) |
|  | Yes | 28 (65.1) | 38 (40.0) | 0.0107 | 66 (47.8) | 88 (62.9) |
| Diabetes | No | 28 (65.1) | 68 (71.6) |  | 96 (69.6) | 134 (95.7) |
|  | Yes | 15 (34.9) | 27 (28.4) | 0.5724 | 42 (30.4) | 6 (4.3) |
| CVD | No | 13 (30.2) | 48 (50.5) |  | 61 (44.2) | 107 (89.9) |
|  | Yes | 30 (69.8) | 47 (49.5) | 0.0415 | 77 (55.8) | 12 (10.1) |
| Astma | No | 37 (86.0) | 83 (87.4) |  | 120 (87.0) | 104 (87.4) |
|  | Yes | 6 (14.0) | 12 (12.6) | 1 | 18 (13.0) | 15 (12.6) |
| Astma/COPD | No | 32 (74.4) | 78 (82.1) |  | 110 (79.7) | 100 (84.0) |
|  | Yes | 11 (25.6) | 17 (17.9) | 0.4171 | 28 (20.3) | 19 (16.0) |
| Dp-ucMGP<br>(pmol/L) | Median<br>(IQR) | 1368<br>(889; 2340) | 771<br>(590; 1174) | 0.0002 | 889<br>(646; 1441) | 508<br>(420; 592) |
| Dp-ucMGP<br>(pmol/L) | Mean<br>(95% CI) | 1445<br>(1148; 1820) | 877<br>(778; 995) | <.0001 | 1022<br>(912; 1151) | 509<br>(485; 540) |

BMI: body mass index; CVD: cardiovascular disease; COPD: chronic obstructive pulmonary disease; IQR: Inter Quartile Range; SD: standard deviation; Dp-ucMGP: dephosphorylated and uncarboxylated Matrix Gla Protein

**Figure 1:**
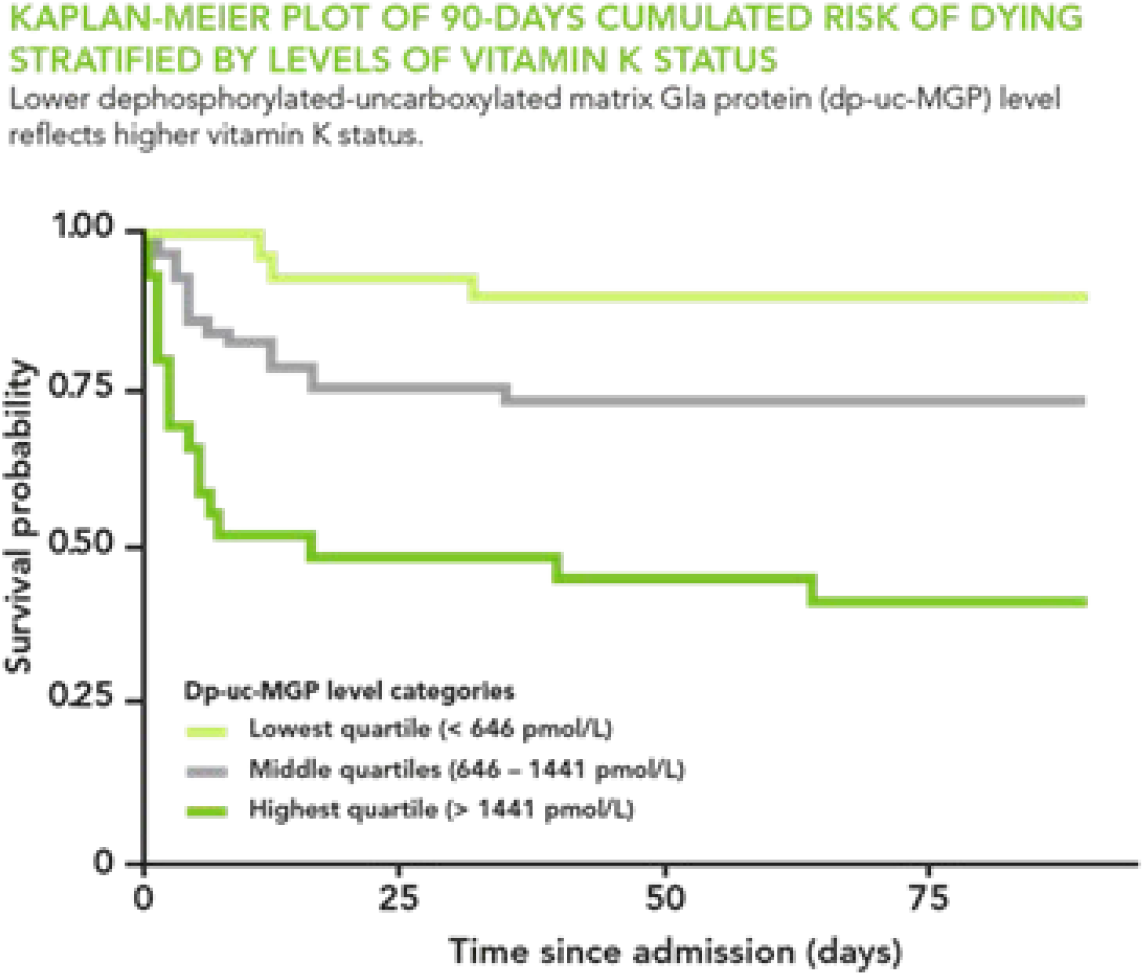
Kaplan-Meier plot of 90-days cumulated risk of dying versus time from blood drawing stratified by levels of dephosphorylated uncarboxylated Matrix Gla Protein (dp-ucMGP) categories (lowest quartile, the two middle quartiles, and highest quartile of dp-ucMGP). High levels of dp-ucMGP reflects low vitamin K status. P-value for log-rank test for comparison of the groups was <0.0001.

A correlation matrix of a selection of measured blood biomarkers and clinical parameters is presented in Figure 2. Dp-ucMGP was positively associated with plasma creatinine (0.56, P<0.0001) and urea (0.58, P<0.0001) and inversely with alanine aminotransferase (−0.25, P=0.01) and lactate dehydrogenase (−0.33, P=0.002). All other correlations were statistically insignificant.

**Figure 2:**
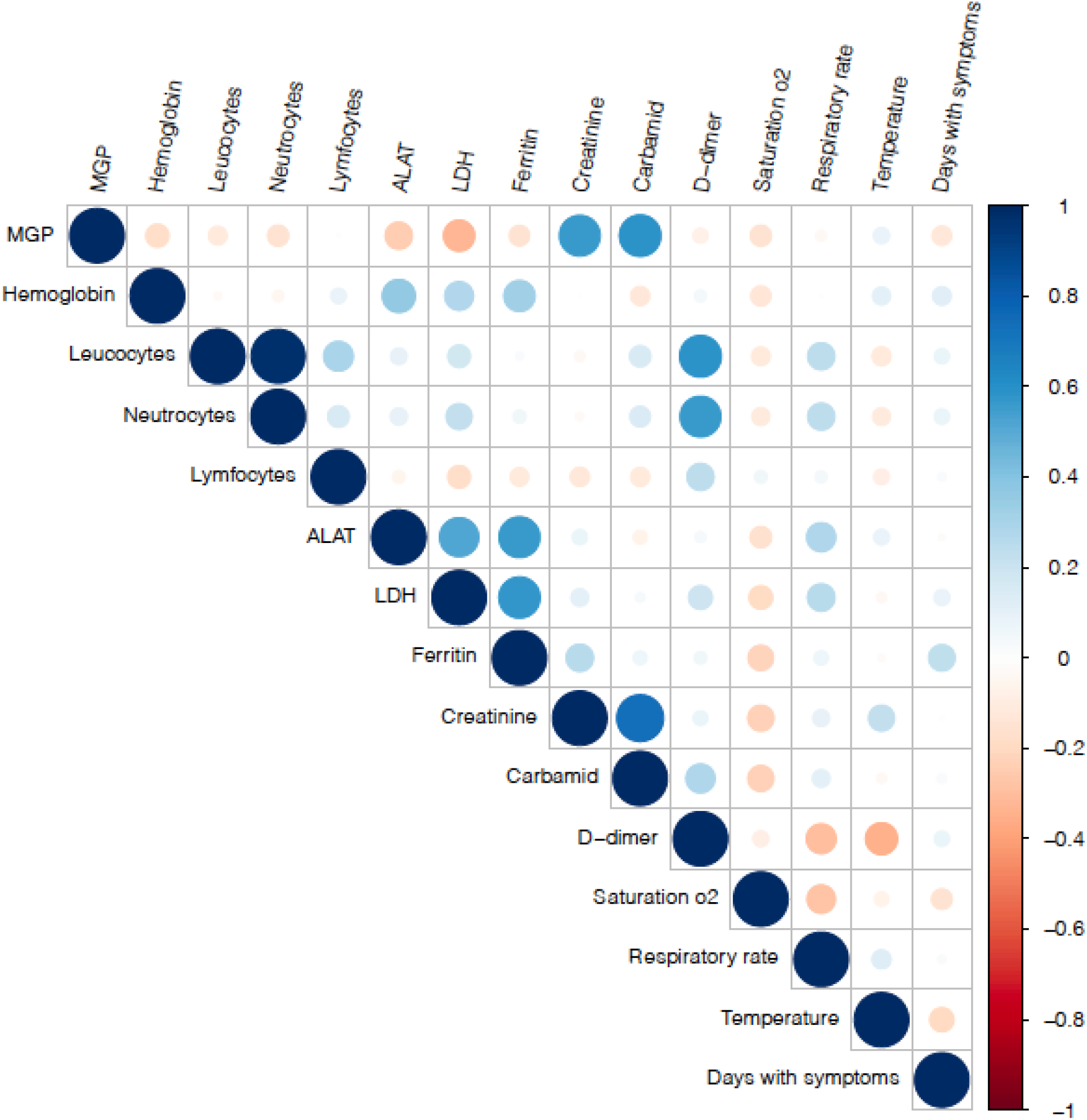
A correlation matrix of measured blood biomarkers and clinical parameters in hospitalized COVID-19 patients. Blue and red colored circles represent positive and negative correlations, respectively. The size of circles reflects the strength of the correlations.

## 4. Discussion

Our results showed that vitamin K status is lower in hospitalized patients with COVID-19 compared to population controls and that low vitamin K status predicts higher mortality among patients with COVID-19. These findings suggest that vitamin K could play a role in the disease mechanisms in COVID-19.

SARS-CoV-2 infection may lead to hypercoagulability and thrombosis,^14,15^ also in some patients in high-dose anticoagulative treatment^16^. The pattern of thrombotic events appears to be different compared with severe pneumonia caused by influenza.^17^ It may appear somewhat surprising that COVID-19 patients, who seem prone to thromboembolism, have lower Vitamin K status, since lowering Vitamin K function pharmaceutically by Vitamin K antagonists is commonly used as antithrombotic treatment and prevention of thromboembolic events in high-risk patients. A possible explanation for this apparent paradox may be that during a state of severe Vitamin K deficiency the intrahepatic Vitamin K-dependent carboxylation (activation) of prothrombotic proteins is prioritized, or preserved, on the expense of peripheral activation of Vitamin K-dependent proteins such as the antithrombotic Protein S and the calcification inhibitory MGP. This is supported by the earlier finding of a preserved intrahepatic prothrombotic effect, as reflected by normal levels of PIVKAII, in COVID-19 patients in spite of significantly increased levels of dp-ucMGP.^4^ In addition, the decreased activation of the calcification inhibitory MGP may increase calcification and subsequent degradation of elastic fibres in the extracellular matrix of lung tissue leading to more severe lung damage in COVID-19 patients.

As countries world-wide are experiencing a second or even third wave of the COVID-19 pandemic, there is an urgent need for measures to improve the outcome and long-term consequences of COVID-19. Supplementation with Vitamin K represents an inexpensive and simple-to-use intervention. It is of potential interest that obesity is a predictor of poor outcome of SARS-CoV-2 infection.^18^ This could be in line with our recent report that obesity was strongly associated with higher levels of dp-ucMGP (indicating low vitamin K status) providing a possible explanation for the link between obesity and COVID-19.^2^ There is limited knowledge about the correlation between intake of Vitamin K and morbidity of SARS-CoV-2 infection at population level. However, it has been reported that areas of Japan with a tradition for consumption of high Vitamin K containing foods such as ‘natto’, soy beans fermented by the Vitamin K producing bacteria *Bacillus subtilis*, are apparently less affected by the COVID-19 pandemic.^19,20^

There are several limitations of the present study. Firstly, the observational study design does not allow us to draw definite conclusions regarding causality. Randomized trials are needed to document potential beneficial effects of vitamin K supplementation on the course of COVID-19 disease. Second, we did not have data on Vitamin D status such as blood levels of 25-OH Vitamin D. This is a limitation because it has been hypothesized that Vitamin D and K could interact in COVID-19 disease.^9^ Third, assessment of lung damage such as CT-scans would have added valuable information on the tissue-specific effects of Vitamin K deficiency during COVID-19. Finally, a long-term follow-up of persistent symptoms among survivors of COVID-19 would have been of great interest to investigate, since any measure that prevents short-term outcomes may also have an influence on long-term outcomes such as persistence of symptoms.

In conclusion, in the present study we confirmed that Vitamin K status is markedly lower in hospitalized COVID-19 patients compared to population controls and that low Vitamin K status predicts mortality in patients with COVID-19. Whether Vitamin K supplementation in COVID-19 patients can change the course of disease and prevent death or long-term consequences of COVID-19 remains to be tested in randomized clinical trials.

## Data Availability

Data are protected by Danish Data Protection Regulations. Requests for data access can be made to the corresponding author, but must be approved by Danish data protection authorities.

## Acknowledgments

The dp-ucMGP measurements were performed and quality assured by Jens Romlund Halgreen and Britt Lisette Corfixen at the Department of Clinical Biochemistry, Rigshospitalet, Glostrup, Denmark

## Author Contributions

Conceptualization, A.L.; Data Curation and Formal Analysis, H.S.; Funding Acquisition, A.L. and T.B.; Methodology and Investigation, A.L., S.B.I., L.R.A., H.L.J., and T.B.; Project administration, A.L.; Resources, C.L., R.W.T., E.A.F., S.L.C. and A.-L.M.H.; Software, H.S.; Supervision, A.L. and T.B.; Writing-Original Draft Preparation, A.L., F.B.K., and S.M.T.; Writing-Review & Editing, A.L., F.B.K., S.B.I., L.R.A., H.L.J., H.S., N.R.J., S.M.T., T.B. All authors read and approved the final manuscript.

